# Extensive antagonistic variants across human genome

**DOI:** 10.1101/2024.11.28.24318135

**Authors:** Beilei Bian, Valentin Hivert, Naomi R Wray, Allan F McRae

## Abstract

Pleiotropic conflict, where a genetic locus has antagonistic effects on different traits, is a common phenomenon observed in animals and plants. While pleiotropy has been widely reported in humans, there is no systematic study of pleiotropic conflict in humans. Here, we leverage GWAS summary statistics of complex diseases and traits derived from large-scale population cohorts to identify pleiotropic regions with conflicting effects. Through a multi-trait colocalization approach, we identified 219 independent regions containing variants showing pleiotropic conflict, which cover ∼11.4% of linkage disequilibrium blocks in the human genome. Antagonistic variants are observed to be enriched for SNPs with intermediate minor allele frequencies and antagonistic regions show signatures of positive/balancing selection. Our results suggest that antagonistic variants are pervasive in humans and indicate their role in maintaining phenotypic and genetic diversity in humans.

## Main

Pleiotropy is defined as a single genetic unit (either a variant or a gene) that influences multiple traits. It is a pervasive phenomenon observed in many species^1-3^. In humans, pleiotropy is observed as a characteristic of many Mendelian diseases^4,5^, and more recently, of human complex traits^6,7^, demonstrating that the human genome is highly pleiotropic^7,8^. For example, sickle cell anaemia is caused by a mutation in the *HBB* gene in chromosome 11, and the abnormal red blood cells further influence other tissue functions, including the eyes, liver and heart^9^. A survey of GWAS-associated loci showed 90% of GWAS-associated loci overlap with more than one trait^7^. The extent of pleiotropy in the human genome is further highlighted by extensive genetic correlations observed between pairs of traits^7,10^.

While most pleiotropic loci have concordant effects for disease risk, a subset of pleiotropic loci have opposing effects on human diseases, resulting in pleiotropic conflict. Such conflict has been reported in several empirical studies of human diseases. For example, discordant genetic effects have been reported across brain disorders^11^, with a meta-analysis across multiple disorders identifying three loci that have opposite effects on schizophrenia and depression, and two between schizophrenia and autism^11^. The shared genetic variants associated with immune-mediated diseases often have opposite effects, with a risk variant for one disease being protective for another^12,13^. Interestingly, many opposite directions of genetic effects on gene expression (eQTLs) have been identified across tissues in humans^14^, suggesting a potential mechanistic explanation for the discordant pleiotropy observed between diseases.

Antagonistic pleiotropy (AP), where a genetic variant controls a trait that is beneficial to an organism early in life but also controls another trait that is detrimental later in life, is the most well-studied example of pleiotropic conflict, with examples seen in humans. For instance, as a tumour suppressor, p53 can prevent cancer by interrupting abnormal cell proliferation, but the increased activity of p53 can simultaneously contribute to ageing^15^. Coronary artery disease (CAD) risk loci have been identified to be antagonistically related to reproductive success^16^. A comparison of genetic variants associated with diseases that occur at different life stages supported the role of AP in ageing^17^.

While the study of pleiotropy in humans is limited experimentally, the increasing availability of biobank cohorts and population-based disease collection for GWAS^18,19^ provides an avenue to assess the extent of such conflict empirically. Here, we use GWAS summary statistics to identify antagonistic regions in the human genome through multi-trait colocalization analysis. We demonstrate extensive pleiotropic conflict both within and across trait biological domains (International Statistical Classification of Diseases 10th Revision codes). We observe signatures of positive and balancing selection at or nearby the identified antagonistic regions.

## Results

### Ubiquitous pleiotropic conflict across human complex traits

The overview of the study is shown in **Fig. 1**. Regions of pleiotropic association were identified using GWAS summary statistics from FinnGen R6, UK Biobank and other large GWAS studies using individuals of European genetic ancestry (inferred from genetic principal components), including binary and quantitative traits (**Supplementary Table 1**). Each quantitative trait was linked to human fitness using the evidence of natural selection observed with modern traits in a previous study^20^ to provide a direction of effect to enable comparison with disease traits. For example, higher heel bone density, early age at first birth and later menopause were assumed to be linked to higher fitness. We took a systematic approach to identify pleiotropic regions and pleiotropic SNPs. We define a sentinel association as the most associated SNP in a chromosomal region. It is well recognised that GWAS sentinel SNPs do not necessarily represent causal variants, but they are expected to be correlated to causal variants. Hence, a pleiotropic causal variant could be tagged by different sentinel SNPs in different GWAS. So, for each GWAS study, we first performed clumping to identify a set of roughly independent trait-associated sentinel SNPs (*P* < 1 × 10^-7^, *r*^2^ < 0.1). The MHC region is the most pleiotropic region in the genome. Multiple studies have reported the pleiotropic effect of classical HLA alleles across phenotypes^21-23^. Given that this region has been well studied and the existence of long LD, the MHC region (chr6:24Mb∼34Mb) was excluded from downstream analysis. Rare variants (MAF < 0.01) were also removed from our study. Potential pleiotropic regions were identified by considering a 2Mb region centred around each sentinel SNP across all traits, followed by colocalization analysis on regions associated with two or more traits. SNPs prioritized by colocalization analysis having opposite effects on different traits were considered candidate antagonistic SNPs. Here, we report those loci showing high regional posterior probability, suggesting it is highly likely that the same associated region is shared by different traits. In other words, we included both high linkage disequilibrium (LD) regions, in which a putative causal variant cannot be pinpointed by colocalization analysis precisely, and regions where traits share the single causal variant with high posterior probability. With this approach, we identified 219 independent loci (194 LD blocks) that discordantly affect different traits and diseases, covering approximately 11.4% of the independent LD regions in the human genome (assuming that there are 1703 independent LD blocks in the European genetic ancestry^24^) (**Supplementary Table 2)**.

**Fig. 1.**
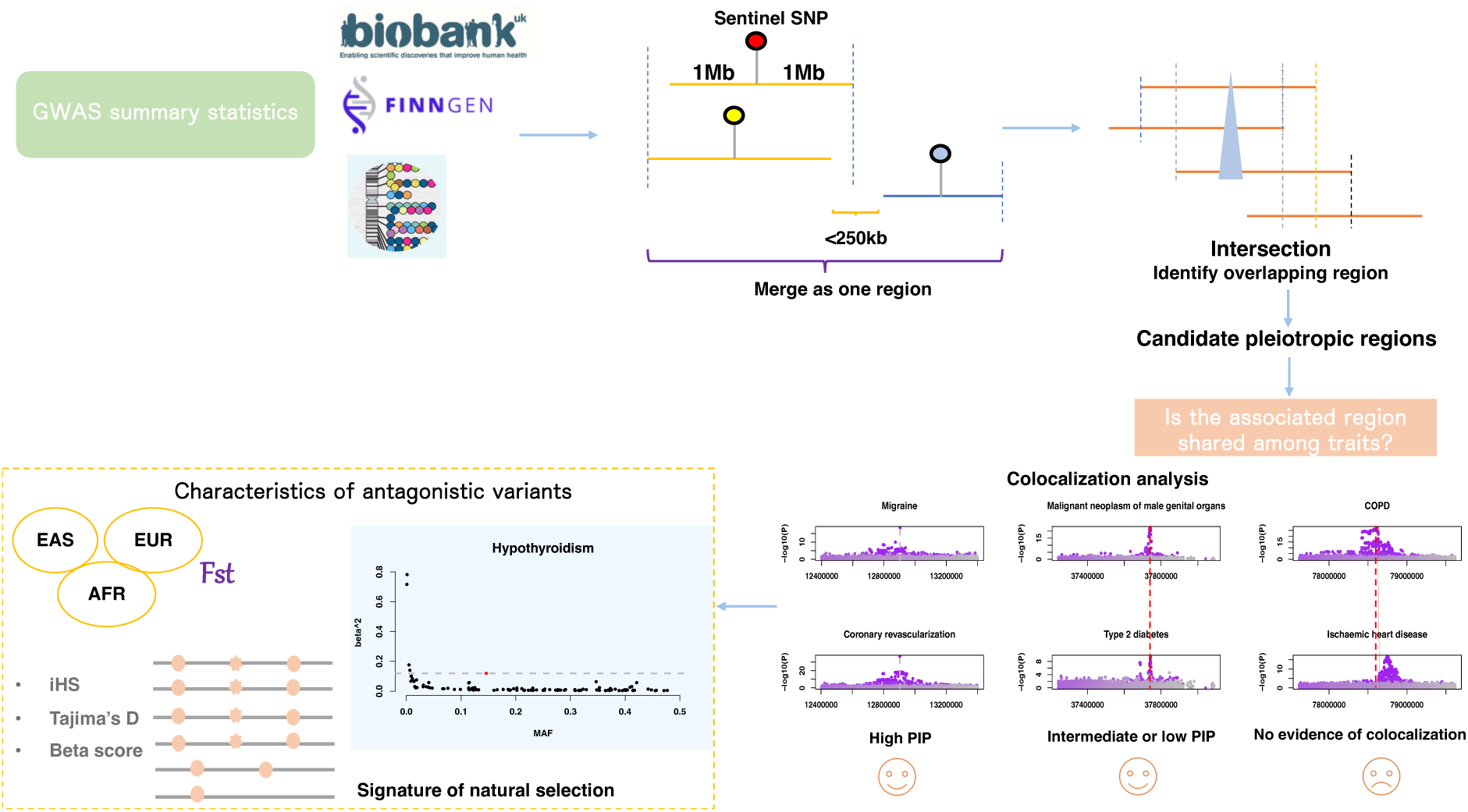
The overview of the study. GWAS summary statistics were collected from UK Biobank (Neale lab round 2 summary statistics), FinnGen R6 and GWAS catalogue. Sentinel SNPs for each GWAS study were defined with an LD reference matched to the population. For each GWAS, an associated region was defined as 1Mb nearby the sentinel SNP. Overlapped associated regions and associated regions close to each other (< 250kb) were merged into one region. Pleiotropic regions were identified by intersecting associated regions of all traits. This was followed by colocalization analysis. Finally, we investigated the characteristics of antagonistic variants (eg., minor allele frequency and effect size) as well as the signatures of selection on each region.

To aid the discussion of results, we have broadly classified diseases as autoimmune diseases and non-autoimmune diseases. For non-autoimmune diseases, ICD-10 was used to assign each disease to a biological domain (for example, respiratory system, **Supplementary Table 3**). We observed that most of the identified loci (211 of 219) affect traits from different biological domains, suggesting that these genomic regions have effects on a wide range of phenotypes. Pleiotropic conflicts were identified within and across biological domains, with malignant neoplasm (77), autoimmune (68), circulatory (66), endocrine diseases (62) and digestive diseases (60) having the largest number of conflicts with other diseases within and across domains (**Fig. 2a**). Autoimmune, circulatory, respiratory, endocrine, metabolic, nutritional diseases and cancers have a higher rate of conflict with diseases in other domains (*P* = 2.96 × 10^-18^, *P* = 3.71 × 10^-9^, *P* = 8.04 × 10^-7^, *P* = 8.37 × 10^-6^, *P* = 1.06 × 10^-2^, *P* = 3.64 × 10^-2^ *and P* = 3.91 × 10^-2^, respectively, one-sided Fisher’s exact test). Conflicts occurred more frequently in diseases from different domains (196 out of 219 (∼90%)) than within domains (23 out of 219 (∼10%)). Genetic variants discordantly affect traits in the same domain were observed across autoimmune diseases (9 loci), female reproductive traits (5 loci), diseases of the digestive system (3 loci), diseases of the nervous system (3 loci) and others. Many conflicts were identified across domains. For example, we observed that 16 loci are shared between autoimmune diseases and endocrine diseases (11 occur between thyroid diseases with and without immune component), and 11 loci are shared between autoimmune diseases and respiratory diseases. Diseases of the circulatory and digestive systems shared 10 antagonistic loci. Migraine and cardiovascular diseases, which partly share genetic etiology^25^, were observed to be discordantly affected by 4 genetic loci. Additionally, some genetic loci were identified to be discordantly associated with the risk of cardiovascular diseases, lipid metabolic disorders and disorders of the gallbladder. Inverse associations were also observed between type 2 diabetes and malignant neoplasms such as breast and prostate cancer. Compared with SNPs showing non-antagonistic associations with multiple traits, the antagonistic SNPs were found to be associated with traits across more biological domains (*P* = 5 × 10^-4^, Fisher’s exact test) (**Fig. 2b**).

**Fig. 2.**
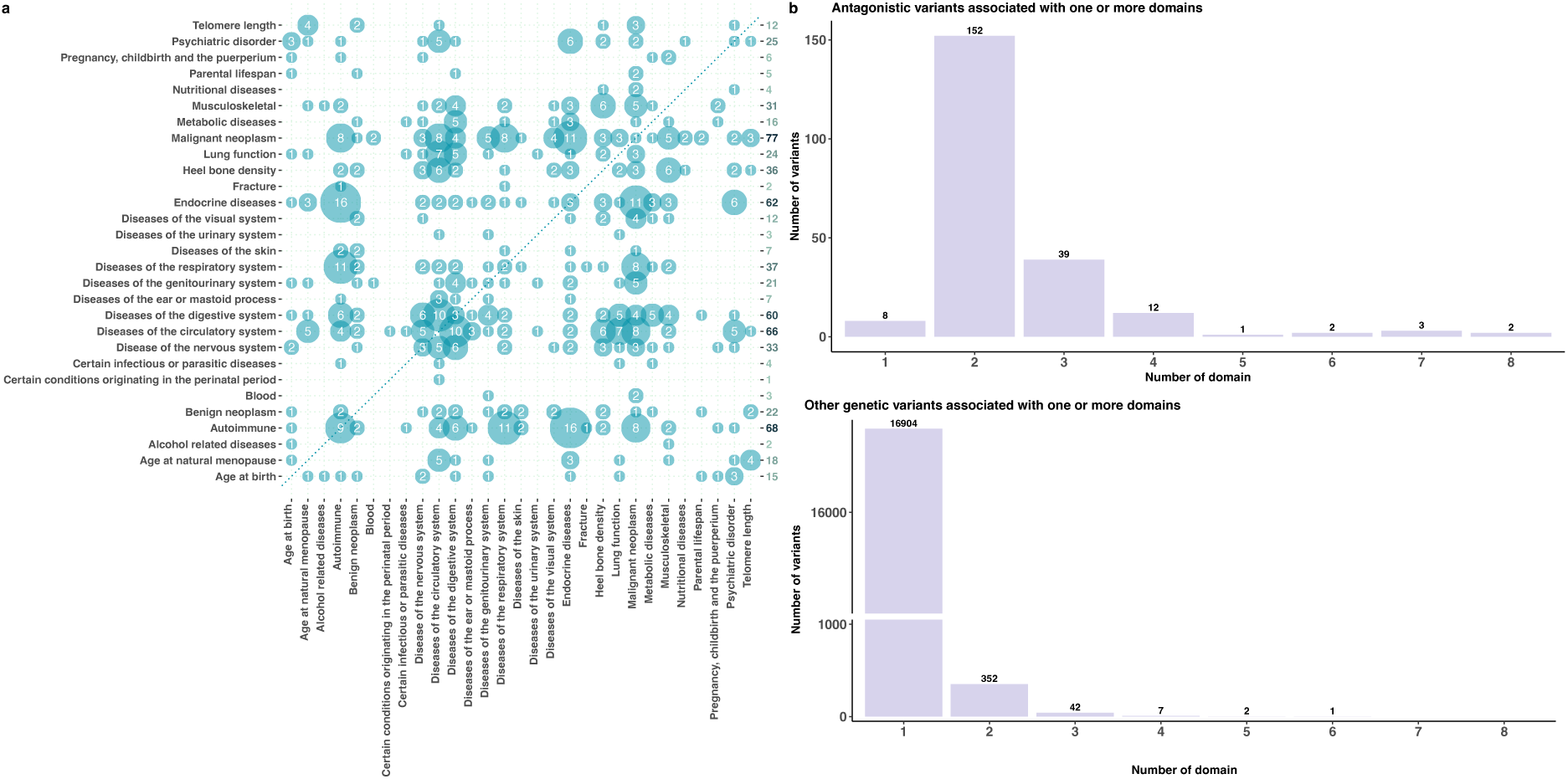
Antagonistic variants affect traits within and across biological domains. (**a**) The number of pleiotropic conflict events across each domain pair. Different domains are given along the x-axis and y-axis. The number in each bubble indicates the number of conflicts identified in each domain pair. The total number of identified conflicts for each biological domain is given along the y-axis (right). (Blood: Diseases of the blood and blood-forming organs and certain disorders involving the immune mechanism) (**b**) Count of genetic variants associated with diseases/traits from different numbers of domains for antagonistic variants and non-antagonistic variants.

### Replication in Biobank Japan

To investigate whether the identified pleiotropic conflicts are shared among ancestries, we looked up the 219 identified antagonistic variants in Biobank Japan. Three pairs of associations were replicated at the level of genome-wide significance. These include *GCKR*, which discordantly affects type 2 diabetes and cholesterol metabolic disorder; *PSCA*, a locus that has an opposite effect on gastric ulcer and gastric cancer; *HNF1B*, the associated variants in this locus, have an opposing effect on the risk of prostate cancer and type 2 diabetes. We identified 62 trait pairs with at least one trait that can be found in Biobank Japan for each direction. Of these, eleven (17.7%) antagonistic variants were replicated at the intermediate significance of 8.06 × 10^-4^ (0.05/62) (**Supplementary Table 4**).

### Gene sets and gene ontology analyses imply immune and cancer-related pathways

Putative causal genes for the identified pleiotropic conflict loci were linked using the locus-to-gene and variant-to-gene models provided by the Open Targets Genetics^26^ (**Supplementary Table 5**). Open Targets Genetics leveraged functional data and QTLs data to prioritize causal genes and identified a total of 354 putative causal genes for the 219 investigated regions (160 regions were linked to a single gene). Gene-set enrichment analysis was performed on the putative causal genes by FUMA^27^, which highlighted 24 gene sets (**Supplementary Fig. 1a**). These are mainly related to immune, cancer, cell proliferation and ageing. Specifically, ten genes highlighted by FUMA are involved in TNF alpha signalling via NF-KB, an inflammatory response pathway regulated by NF-KB. Other immune-related pathways are also implicated, such as interferon-gamma response, IL-2-STAT5, and complement. The enrichment in the p53 pathway is driven by six genes. These genes are particularly involved in DNA maintenance, controlling cell proliferation and cell cycle. The IL6/JAK/STAT3 signalling pathway, which is involved in both immune responses and cancer, was also highlighted. In terms of molecular function, we observed that these genes are strongly enriched in the DNA binding pathway, suggesting their roles in gene regulation (**Supplementary Fig. 1b**). Notably, of the 354 linked genes, 58 were linked to transcription factor activity (P_adjusted_ = 1.76e-14).

### Antagonistic SNPs have a higher minor allele frequency than SNPs that have concordant effects

We compared the minor allele frequency distribution of the putatively causal antagonistic SNPs (**Supplementary Table 2**) to SNPs that have concordant effects (**Supplementary Table 6**) on different traits from more than one biological domain. Different MAF distributions were observed between these two groups (Kolmogorov–Smirnov test *P* = 2.78 × 10^-4^, **Fig. 3a**). For antagonistic variants, the proportion of variants in each MAF bin increased with MAF. In contrast, variants that have concordant effects on traits tend to be uniformly distributed in different MAF bins. However, the detection power is determined by allele frequency of risk allele, genotype relative risk, number of cases and controls, significance threshold and lifetime risk of disease. For antagonistic variants detection, the risk minor alleles will have more power to be detected than the risk major alleles, assuming all the other parameters are the same constant for different diseases. Given that the GWAS summary statistics were collected from large biobanks and consortiums, we expected ascertainment bias in our comparison to be limited.

**Fig. 3.**
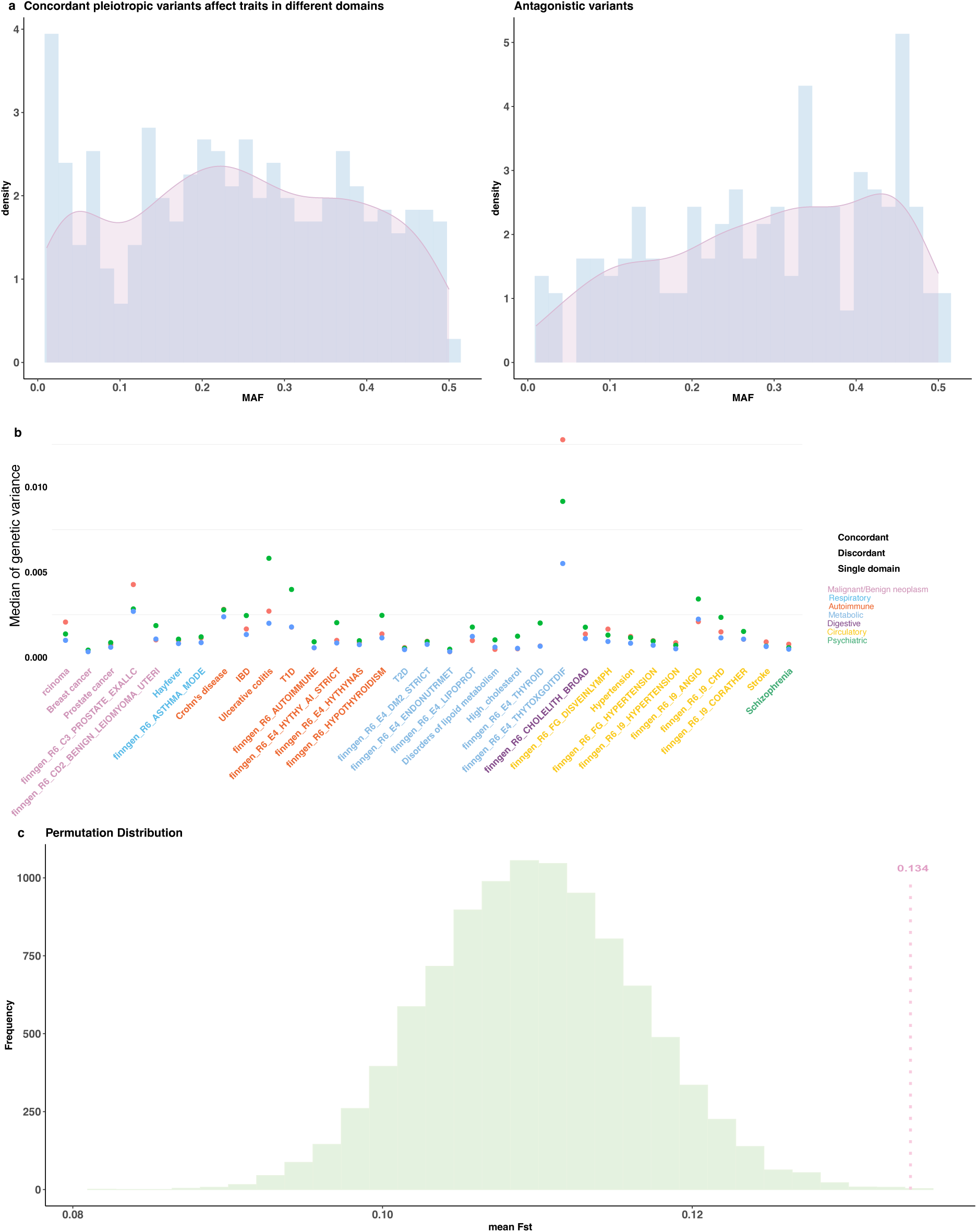
Characteristics of antagonistic variants and the comparison between antagonistic variants and non-antagonistic variants. (**a**) Comparison of minor allele frequency probability density distribution between SNPs that have concordant effect on traits in different domains and antagonistic SNPs. The left figure is the allele frequency distribution of putatively causal SNPs that have concordant effects on traits in different domains. The right figure is the allele frequency distribution of putatively antagonistic causal SNPs. (**b**) Comparison of genetic variance explained by antagonistic, concordant and single domain SNPs. Each dot represents the median value of genetic variance explained by SNPs associated with the given trait. (**c**) The differences between the mean value of observed *F*_*st*_ and permutations. The mean value of *F*_*st*_ is given along the x-axis. Y-axis represents the frequency of occurrences over a range of mean *F*_*st*_ in 10,000 permutations. The green bars show permutation distribution. The vertical pink line is the mean value of the observed *F*_*st*_.

We also compared the genetic variance explained by (1) antagonistic SNPs, (2) SNPs with concordant effect on traits from different domains and (3) SNPs that affect traits in the same domain. Across a range of traits with large numbers of GWAS associations, the genetic variance of antagonistic SNPs was on average higher than those SNPs showing concordant effects across domains or SNPs affecting traits in a single domain, explaining the largest genetic variance in 21/33 traits (**Fig. 3b**).

### Antagonistic SNPs exhibiting a divergent natural selection

Considering the identified antagonistic SNPs were enriched at intermediate allele frequency, we then asked whether these regions were under selective pressure. We first calculated Wright’s fixation index *F*_*ST*_ of each SNP using three populations (EUR, AFR and EAS) from 1KG data. Briefly, *F*_*ST*_ is a means of assessing population structure and an estimate of the variance of allele frequency among populations. This indicator reflects multiple evolutionary processes, including selection, migration, mutation and genetic drift^28^. It has been commonly used for identifying signatures of divergent selection, with loci exhibiting larger *F*_*ST*_ being candidates that are subject to natural selection^29,30^. Here, we generated 10,000 sets of matched control SNPs by sampling SNPs from the 1KG EUR data, with MAF and LD scores matched to the observed antagonistic SNPs. We then sampled each control SNP from its matched control list and calculated the average *F*_*ST*_ among the control SNPs. We observed strong evidence that the observed antagonistic SNPs have larger *F*_*ST*_ compared to the control SNPs and pleiotropic SNPs that have a concordant effect (**Fig. 3c**).

### Signatures of natural selection in antagonistic regions

The larger effect size, MAF and *F*_*ST*_ of antagonistic variants indicate that some antagonistic regions are putatively under natural selection. We further used other population genetics metrics, including the integrated haplotype score (iHS)^31^, Tajima’s D^32^ and *β*^(2)^ statistics^33^ to detect signatures of natural selection in antagonistic regions. In brief, iHS is a metric that quantifies the signature of positive selection using haplotype data. A larger |iHS| score indicates a signal of positive selection with an unusually long haplotype. Tajima’s D is a commonly used test of neutrality. *β*^(2)^ score is used to test long-term balancing selection using allele frequency correlation. These analyses were performed on UK10K unrelated individuals. For each antagonistic SNP, we extracted iHS and *β*^(2)^ values of SNPs in LD (*r*^2^ > 0.3) with it (**Supplementary Tables 7 and 8**) and calculated Tajima’s D around the identified antagonistic SNPs (**Supplementary Table 9**). The empirical distribution of each metric was generated from the whole genome results. We observed that 35 (16.5%) of the tested 212 regions (MAF of antagonistic variants greater than 0.05) have |iHS| greater than the 1% threshold of the empirical distribution (|iHS| = 2.6), while 17 (7.8%) regions have a *β*^(2)^ greater than the 1% threshold of the empirical distribution (*β*^(2)^=9.43). The whole genome distributions of |iHS|, Tajima’s D and *β*^(2)^ are shown in **Supplementary Figs. 2-4**.

#### Immune-related loci have opposite effects on different traits showing signatures of natural selection

We identified several antagonistic variants associated with diseases regulated by the immune system that are under selection, suggesting that the genetic trade-off might be due to the adaptation to pathogens in the past. For example, we identified signatures of positive selection at the *PTPN22* locus, which regulates T-cell activities. The SNP rs2636014 that was identified to be under positive selection (|iHS|=3.12) is in weak LD with rs2476601 (*r*^2^ = 0.34), a missense variant discordantly affecting autoimmune diseases and basal cell carcinoma (**Fig. 4a**). Similarly, the TLR cluster was also detected with rs10008032 (|iHS|=3.6) in the same LD region with rs5743618 (*r*^2^ = 0.34), a missense variant in the innate immune gene *TLR1* (**Supplementary Fig. 5a**). This SNP was identified to be strongly associated with hay fever and breast cancer in opposite direction. Additionally, previously reported loci such as *SH2B3/ATXN2* and *FUT2* also showed a signature of positive selection with the UK10K data (**Supplementary Figs. 5b and c**). We also identified some immune-related loci under balancing selection. A striking example is the *NFKB1* locus, a transcription factor that can be activated by bacteria and viruses and regulates inflammatory responses, showing larger Tajima’s D value (regional largest Tajima’s D = 3.21) and *β*^(2)^ statistics (largest *β*^(2)^ = 12.95) (**Fig. 4c**). The signature of balancing selection at this locus is colocalized with GWAS signals of hay fever and primary biliary cirrhosis. We also detected that the *CTLA4* locus, which regulates T-cell responses, is under balancing selection (**Fig. 4d**). These suggest that the conflict effects on different diseases might be due to different haplotypes that have been maintained for a long time.

**Fig 4.**
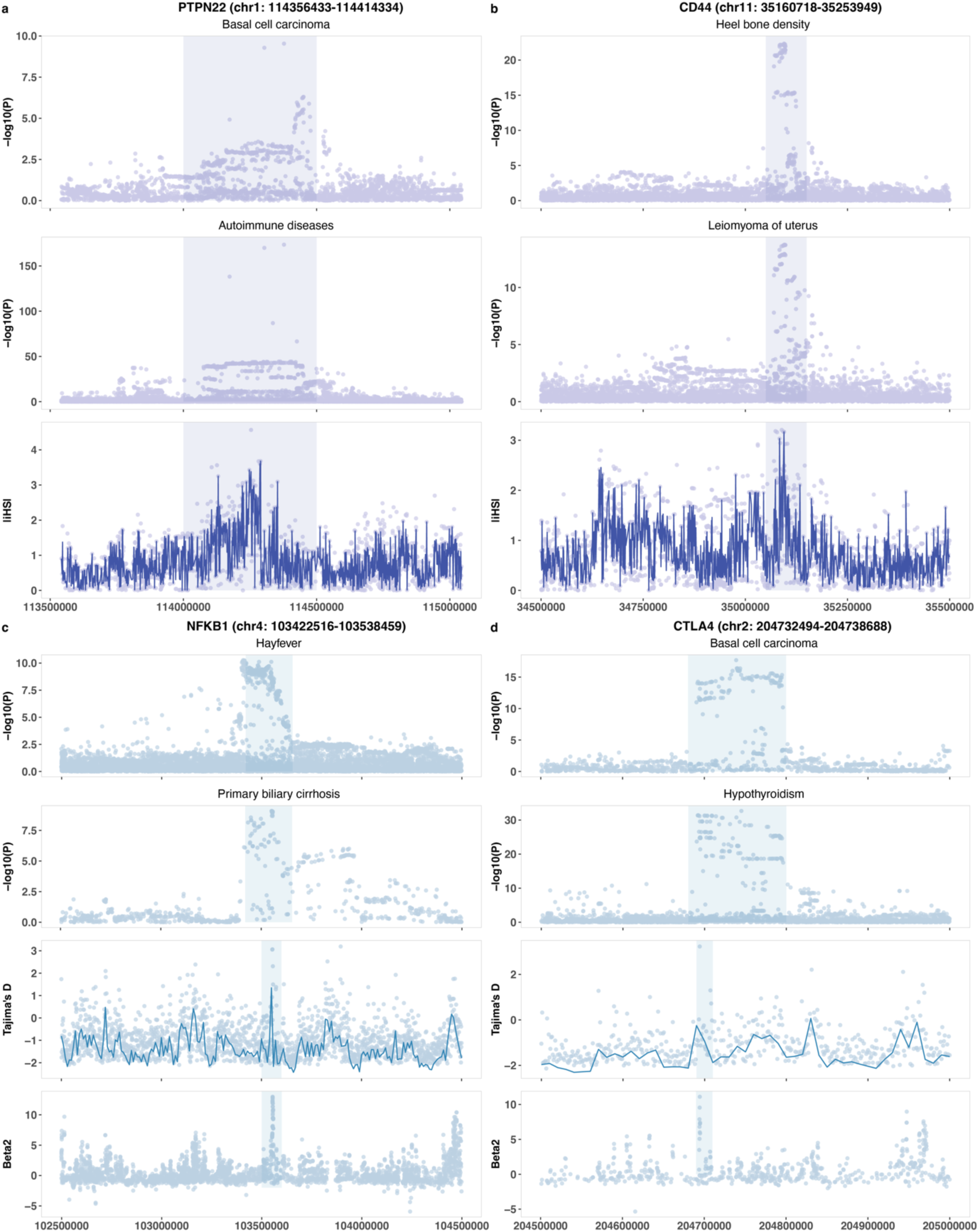
Selected examples of signatures of selection observed in antagonistic regions. The top two figures are GWAS signals of two traits, which are affected by the locus in opposite direction. Each dot represents a SNP. The x-axis is the genomic position, while the y-axis is −*log*_10_(*P*). (**a-b**) The bottom figure shows |iHS| in the genomic region. Each dot indicates |iHS| of a single SNP, while the line connects the average |iHS| within 500bp. **(c-d)** The third row is Tajima’D in the genomic region. Each dot indicates Tajima’s D value calculated with window size of 1kb, while the line connects Tajima’s D value calculated with window size of 10kb. The bottom figure shows beta score in the genomic region. Each dot indicates the beta score of a single SNP. The colocalized GWAS and selection signals are highlighted in rectangle.

#### Essential genes are identified to show signatures of natural selection

In addition to immune-related loci, we detected several essential genes, including transcription factors and other genes that participate in basic activities throughout life. For example, we identified that the *CD44* locus is under positive selection and discordantly affects heel bone density and uteri leiomyoma (**Fig. 4b**). *CD44* gene is involved in cell-cell interaction, cell adhesion and migration, and is highly expressed in many cancers^34^. It was also identified for bone loss^35^. Another example is the *BNC2* locus, which encodes a conserved zinc finger protein and has opposite effects on age at first birth and lifespan. Some transcription factors and genes regulating gene expression showed signatures of balancing selection. These include the *NFKB1* gene, *BICC1* gene and *SOX9* gene (**Fig. 4c, Supplementary Figs. 6a and b**). The *BICC1* gene encodes an RNA-binding protein and plays a fundamental role in regulating gene expression, which is associated with heel bone density and glaucoma in the opposite direction.

#### Other notable genes under natural selection

Our results also showed some other interesting examples, including the *MAPT/CRHR1* cluster, *PSCA*, *INHBB*, *CFDP1*, *TSPAN10* and *ADCY3* genes. We observed a signature of positive selection at the *INHBB* locus (**Supplementary Fig. 5d**), which encodes a subunit of both activin and inhibin participating in stimulating and inhibiting FSH secretion. The SNP rs4849879(G) was associated with a lower risk of breast hypertrophy, which is related to hormonal changes and may occur during puberty, but a higher risk of breast cancer. The region containing the *TSPAN10* locus has an opposing effect on the risk of basal cell carcinoma and eye disorder (**Supplementary Fig. 5e**). Interestingly, this locus is strongly associated with hair colour and skin colour. The *CFDP1* locus, which has an opposing effect on cardiovascular diseases and migraine, was identified under positive selection (**Supplementary Fig. 5f**). The *MAPT/CRHR1* cluster was found to be under natural selection^36^ and has been discordantly associated with Parkinson’s disease and neuroticism (**Supplementary Fig. 6c**). Similarly, we observed signature of balancing selection at the *ADCY3* locus, with a putative causal SNP rs4343432(A) associated with a higher risk of breast cancer but lower risk of Crohn’s disease (**Supplementary Fig. 6d**). Another example is that the *PSCA* locus (rs1045547) has an opposite effect on the risk of stomach cancer and duodenal ulcer, which was detected to be under balancing selection^37^ (**Supplementary Fig. 6e**).

### Antagonistic regions are associated with life history traits

We looked at the age of onset of each disease in each trait pair and classified the antagonistic regions into 3 classes, early versus early, early versus late and late versus late. We observed that among the 168 disease trait pairs, 115 (68%) of them are affected by the pleiotropic conflict that occurs only late in life. This is followed by 51 (30%) genetic regions associated with conflict between early-onset and late-onset diseases, consistent with the widely studied antagonistic pleiotropy phenomenon (**Fig. 5a**). Examining regions affecting pairs of traits with onset early and late in life, we observed that one third of the loci are linked to genes related to the immune system, which is consistent with our gene set enrichment results. A subset of examples is shown in **Fig. 5b**. For example, we identified several genetic variants that discordantly affect immune-related diseases and malignant neoplasms. The SNP rs5743618 (A) is associated with decreased risk of asthma and hay fever and is also associated with an increased risk of breast cancer. Another interesting SNP is rs3087234 (A) in the *CTLA4* gene, which encodes a protein on T cells that regulates immune responses, significantly associated with a lower risk of autoimmune diseases, including Type 1 diabetes and myxoedema, but a higher risk of basal cell carcinoma. Similarly, this was also observed in the *TICAM1* gene, which mediates TLR signalling pathways. The SNP rs7254729 (C) was identified to be associated with a lower risk of thyroid autoimmune disease but a higher risk of basal cell carcinoma.

**Fig 5.**
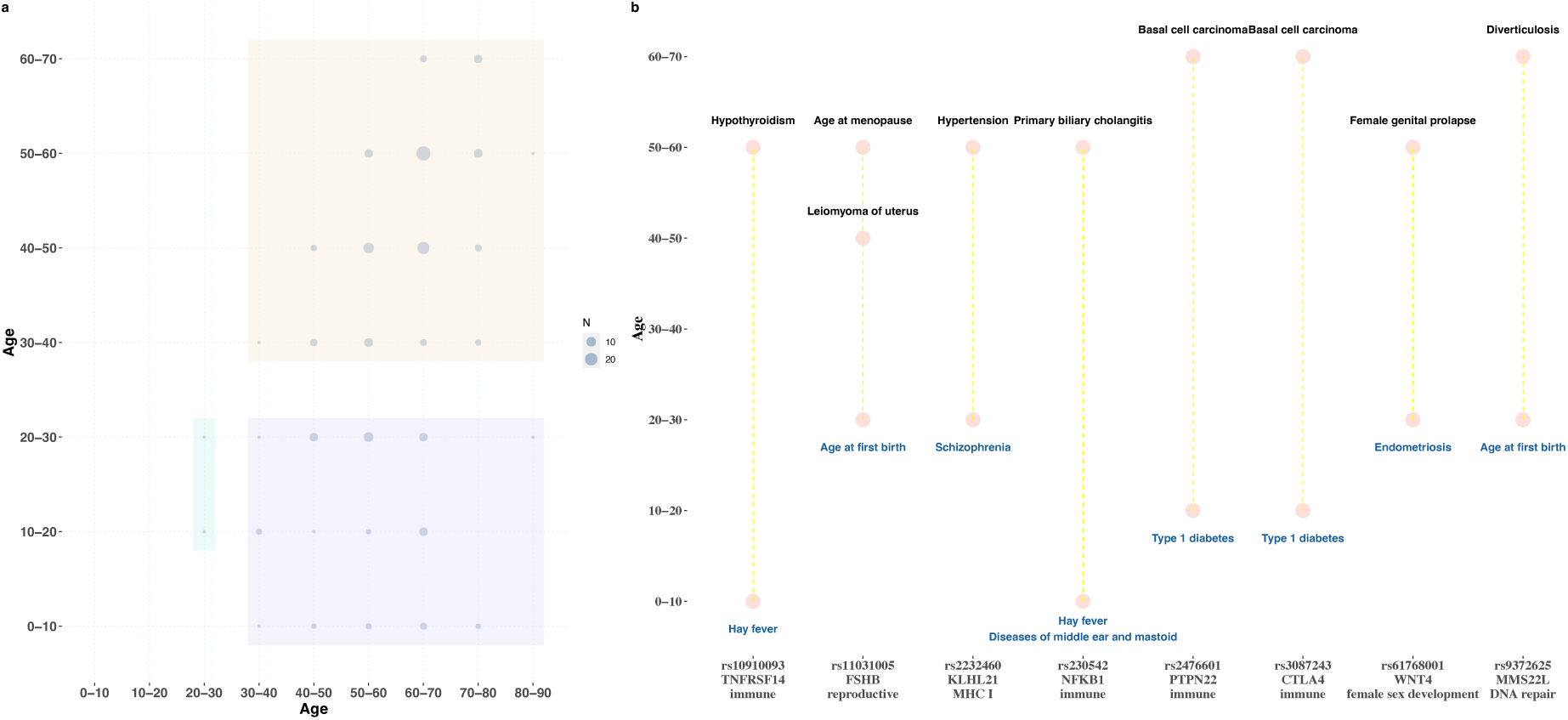
Antagonistic regions affect disease pairs in late-late and early-late age of onset patterns. (a) The number of disease pairs identified at different age groups. The circle size indicates the number of disease pairs in each age group pair. Early-early, early-late and late-late pairs are highlighted in different colors. (b) Selected examples showing antagonistic regions affect life-history traits. Diseases/traits affected by the genomic region in opposite directions are marked in black and blue.

Hormones have been implicated to have conflicting effects and exhibit trade-offs on traits that occur at different life stages^38^. We observed that some genetic variants that play a role in the reproductive system have opposite effects on traits occurring at different life stages. These include: rs11031005 (T), close to the *FSHB* gene and is associated with early age at first birth. However, it is associated with a higher risk of genitourinary system diseases and early age at natural menopause. This suggests genetic variant associated with late reproductive behaviour is beneficial to ovarian ageing, potentially mediated by hormones. Genetic variants at/nearby WNT4 locus were associated with discordant risk of endometriosis and female genital prolapse, which occur at early and late stages of life. The WNT4 protein is responsible for the production of androgens and the development of the female reproductive system. We also identified an intronic SNP rs17601876 (G) in *CYP19A1* that is associated with higher heel bone density and a higher risk of endometrial cancer. *CYP19A1* is involved in the conversion of androgens to estrogen, which is involved in bone growth. Taken together, these indicate that hormone-related genes play an antagonistic role in the early and late stages of life.

## Discussion

Understanding pleiotropic regions having conflicting effects on different diseases is important for human health, especially drug development^39^. Our study has demonstrated that antagonistic variants for human contemporary diseases are widespread throughout the genome. In this study, we leverage GWAS results and multiple population genetic metrics to characterise antagonistic regions and found that putative causal antagonistic SNPs are more differentiated across populations. Many of these genomic regions affect life history traits.

We observed strong evidence for immune-related loci. Some of them affect life history traits (**Fig. 5b**). Previous studies have implicated that early-life infections and inflammations are considered to play an important role in adult diseases^40^. In particular, alleles responsible for infections at the early stage of life were found to have opposite effects on longevity and adult diseases^41^. This conflict might be due to evolutionary forces, such as positive selection and balancing selection, which explains the high proportion of common variants observed in this study. Studies that analysed ancient DNA data showed that adapted alleles in the past contribute to the risk of autoimmune diseases today^42^.

Our disease age of onset analysis showed that a large proportion of pleiotropic conflict variants occur between late-onset diseases and between early and late-onset diseases. Interestingly, we observed loci related to hormones have conflicting effects on life-history traits, consistent with the fact that hormones regulate physiological function at different time points. In addition, we observed that putative causal genes are highly enriched in transcription activities. A study examining pleiotropy at the cellular network level shows that the pleiotropic gene modules are enriched for basic physiological functions^43^, but the direction of the effect of gene modules needs to be investigated in the future.

While our study has provided a comprehensive study of antagonistic variants in the human genome, we are limited by the availability of the GWAS data we used to make these inferences. First, there remains substantial difficulty in the identification of causal variants from GWAS data, so we were only able to identify the shared antagonistic regions. Therefore, whether the opposite effect on target traits was driven by the same variant is unknown. Secondly, limited by the range of diseases examined in GWAS, we could not identify what exactly drives selection. For example, genetic changes due to ancestral exposure to pathogens are not able to be quantified, and we rely on modern-day disease as a proxy. Thus, there are likely many more regions of antagonistic effects in the genome.

Overall, our study provides a picture of widespread antagonistic variants across human diseases, highlighting the complexity of the human genome. We expect the number of identified antagonistic regions to increase as the range of phenotypes examined via GWAS increases, and with biobank scale studies across ancestral diversity. By integrating large GWAS resources and new population genetic methods, we will further understand how genetic and phenotypic diversity is maintained for human complex traits.

## Methods

### GWAS summary statistics

#### FinnGen

We used FinnGen^44^ GWAS summary statistics from release 6, covering 2,861 disease endpoints with a total sample size of 260,405 individuals (**table S1**). Medication-related phenotypes, dental endpoints, injuries endpoints, pregnancy and delivery were not included in the analysis. FinnGen individuals were genotyped with Illumina and Affymetrix chip arrays. The SNP array data were imputed to a Finnish SISu reference panel, which consists of 3,775 whole genomes. The genomic positions of variants are represented by the hg38 assembly.

#### UK Biobank

The GWAS summary statistics of UK Biobank were acquired from Neale Lab (http://www.nealelab.is/uk-biobank) round 2 version. The sample QC and variant QC criteria are described in their documentation (https://github.com/Nealelab/UK_Biobank_GWAS).

We additionally downloaded summary statistics of binary traits carried out by using SAIGE^45^. Some of these GWASs traits overlap with those from FinnGen. We used these as supplement traits to confirm the discovery of antagonistic variants. Briefly, GWASs were conducted on UKB White British participants. All individuals were imputed to the Haplotype Reference Consortium (HRC) panel. The imputed genotype data were downloaded from UK Biobank release 2. The first four principal components, sex and age were included as covariates in the association analysis.

#### Psychiatric Genomics Consortium (PGC)

Summary statistics of psychiatric disorders were downloaded from PGC (https://www.med.unc.edu/pgc/download-results/), including attention-deficit/hyperactivity disorder (ADHD), bipolar disorder (BIP), anorexia nervosa (AN), cannabis use disorder (CUD), autism spectrum disorder (ASD), major depression (MDD) and schizophrenia (SCZ).

#### GWAS Catalogue

We also considered other large-scale GWA Studies, with summary statistics available in the GWAS catalogue. Studies conducted on European individuals were included.

The full list of GWAS summary statistics used in our study is summarized in **Table S1**.

### Defining sentinel SNPs

We first defined sentinel SNPs for each trait. Different LD reference panels were used for these summary statistics according to samples used for conducting GWAS. For FinnGen, the LD estimation was acquired from SISu v3 reference panel as described in FinnGen documentation (https://finngen.gitbook.io/documentation/methods/genotype-imputation/sisu-reference-panel). For UKB studies, we randomly selected 20,000 unrelated individuals from UKB as the LD reference panel. The variants were matched to each study. Specifically, for GWASs carried out by SAIGE, the imputed UK Biobank release 2 data was used as the LD reference panel. For GWASs acquired from Neale lab, the UKB release 3 was used as the LD reference panel. For other GWASs, 1000 Genomes European data was used as the LD reference panel, which consists of 503 individuals. For each trait, we defined the sentinel SNPs with plink1.9^46^ (--clump-kb 500 –clump-p1 1e-7 –clump-r2 0.1).

### The direction of effect on different diseases/traits

We simply defined an antagonistic variant as having a discordant effect on disease traits if the variant increases the risk of one disease but decreases the risk of another disease. For some quantitative traits, we allocated direction according to their relationship with fitness^20^. A reduction in fitness was considered a disease state when defining antagonistic variants with these traits.

### Defining pleiotropic conflict loci

As the terminology “antagonistic pleiotropy” specifically refers to the theory of genetic conflict at different points in the life cycle, we use “pleiotropic conflict” or “antagonistic variants” interchangeably in this study to refer to the wider phenomenon. The genomic positions represented in hg19 were converted to hg38 assembly. We took a 2Mb window around each sentinel SNP, then physically overlapped regions across all traits using BEDTools^47^ (**Fig. 1**). If the genomic region overlaps across traits, then colocalization analysis was performed using HyPrColoc^48^, which allows for a large number of traits to be analysed simultaneously. We extracted a 2Mb genomic region around each sentinel SNP in each trait (1Mb on each side of a sentinel SNP). The overlapped SNPs in a trait set were used in colocalization analysis. Several scenarios were observed: 1) low LD region, high posterior probability and regional probability from colocalization analysis. 2) high LD region, high regional probability, but the shared causal SNP is not distinguishable due to strong LD. 3) different association peaks within the overlapping region, and low regional probability (not colocalized) (**Fig. 1**). Here, we considered the first two scenarios as sharing the associated region. In summary, if the genomic region meets these criteria: 1) it is discordantly associated with different traits, 2) regional probability > 0.8 and posterior probability > 0.5 in colocalization analysis, we consider it a pleiotropic conflict locus.

### Replication of antagonistic variants in Biobank Japan

We took the antagonistic variants identified in the European ancestry study and sought replication in Biobank Japan. We first identified trait pairs that can be potentially replicated (at least one trait in each direction available in Biobank Japan phenotypes). This results in 62 trait pairs with at least one trait in each direction matched to phenotypes available in Biobank Japan. A significance of 8.06 × 10^-4^ (0.05/62) was used for a successful replication.

### Comparison of allele frequency distributions

To compare the allele frequency distributions between pleiotropic variants with concordant effects and antagonistic pleiotropic variants, we included all the putative causal SNPs in pleiotropic regions identified from colocalization analysis. The allele frequencies were calculated using 1KG phase 3 EUR data. We only kept SNPs that are in the 1000G EUR panel. To ensure the variants included in the comparison are independent of each other, we removed variants that are in LD with each other (*r*^2^ > 0.1) and only kept one variant in each LD region. We used 1000G EUR as a reference panel for LD calculation. Finally, the remaining variants were classified into pleiotropic concordant SNPs and discordant SNPs according to the direction of effect on traits.

### Comparison of magnitude of genetic variance

We selected traits/diseases that have at least 3 SNPs in each category: (1) antagonistic SNPs, (2) SNPs with concordant effect on traits from different domains and (3) SNPs that affect traits in the same domain. SNPs in the MHC region were excluded in the analysis. We calculated *f* × (1 − *f*) × *β*^2^ to represent genetic variance.

### *F_st_* permutation test

We tested the *F*_*ST*_ at our pleiotropic conflict SNPs with a matched set sampled from the genome following the procedure described in Guo et al^49^. We first calculated the MAF and LD score of all the SNPs on 1KG EUR data. *F*_*ST*_ was calculated using 1KG EUR, EAS and AFR populations. The *F*_*ST*_ method described in Weir, B.S. 1996^50^ was implemented in GCTA^51^. MAF was set from 0 to 0.5, with an increment of 0.025. In each MAF bin, the LD scores were divided into 20 groups by an increment of 5% percentile. Then each putatively causal antagonistic SNP was allocated into a group of SNPs with MAF and LD scores matched. These groups with antagonistic SNPs allocated were combined as the full control SNP list for the permutation test. Next, for each antagonistic SNP, we sampled one SNP from its matched control group (MAF and LD score matched). For each permutation, we calculated the mean *F*_*ST*_ of the sampled control SNPs. We repeated the sampling procedure 10,000 times. The *F*_*ST*_ distribution of the control SNPs was generated from the permutations.

### Link putative causal variants to causal genes

The causal genes were looked up from the Open Targets Platform^26^, a database covering all the putatively causal genes for published GWAS studies. In general, the causal genes were prioritized by leveraging proximity and functional information. For each associated locus, the causal genes were acquired from the results prioritized by the locus-to-gene (L2G) pipeline. We followed these customized rules to link the shared associated locus to genes: 1) If the locus is not colocalized with a QTL locus, the locus was linked to the gene that shows the highest overall L2G score. 2) If the locus is colocalized with QTL, the locus was linked to the colocalized gene(s) and the gene that shows the highest overall L2G score if these were different. 3) For a locus that is not on Open Target, the prioritized variant was linked to eQTL gene(s) via the variant-to-gene (V2G) pipeline, which was designed by Open Targets Genetics^52^. 4) If it is not an eQTL SNP, it was linked to the nearest gene.

### Gene-set and gene ontology analyses

We take all the linked putative causal antagonistic genes for gene-set enrichment analysis. Gene-set enrichment analysis was conducted using FUMA^27^, with the hallmark gene sets obtained from MsigDB^53^. Gene-set P-value was corrected for multiple testing using Benjamini-Hochberg (FDR) method. Molecular function enrichment was analysed using the GO enrichment analysis tool (http://geneontology.org/). The enriched pathways were reported at *P_FDR_ < 0.05*.

### iHS analysis

To detect recent positive selection, we performed iHS analysis using selscan^54^ on the UK10K data. Briefly, we selected 3647 unrelated individuals using GCTA^51^ (--rel-cut-off 0.05). SNPs with MAF < 0.001, HWE p-value < 10^-6^, and missing genotype call rates > 0.05 were removed. As iHS is a haplotype-based method, which uses phased genetic data as input, statistical phasing was conducted using Shapeit4^55^ with the genetic recombination map downloaded from the 1000 genomes resource (ftp://ftp.1000genomes.ebi.ac.uk/vol1/ftp/technical/working/20130507_omni_recombination_rates/). Genetic variants with MAF < 0.05 were excluded from the iHS analysis. The ancestral state of each SNP was acquired from dbSNP FTP site (https://ftp.ncbi.nlm.nih.gov/snp/organisms/database/shared_data/). In selscan, the unstandardized iHS is calculated as 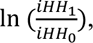 where *iHH*_1_ is the integrated haplotype homozygosity for derived haplotype while *iHH*_0_ is the integrated haplotype homozygosity of the ancestral haplotype. Therefore, a large positive value indicates a long-range haplotype carrying the derived allele; a large negative value indicates a long-range haplotype carrying the ancestral allele. Both positive and negative values were considered in this study since we assumed that selection could act on both ancestral and derived alleles.

### Calculation of Tajima’s D

Tajima’s D in 1kb and 10kb window sizes were also calculated using the UK10K data. VCFtools^56^ was used for the calculation.

### Detection of signals of balancing selection using BetaScan2

To detect signals of balancing selection, we used BetaScan2, which utilizes both polymorphism and substitution data^33^. This can slightly improve detection power compared to Beta1 statistic, which only uses polymorphism data^33,37^. To determine ancestral alleles, we downloaded the epo file, which gives variant states information in the outgroup as the documentation suggested (https://github.com/ksiewert/BetaScan/wiki/Tutorial). This file was also used to call substitutions where the allele of humans differs from the outgroup allele. Then glactools was used to convert UK10K VCF files to ACF files needed by BetaScan. We calculated Beta2 with the divergence time set to 12.5 (in coalescent unit).

### The age of onset of disease traits

For disease traits collected from FinnGen, the information on the age of onset was acquired from https://risteys.finregistry.fi/, which is used for exploring FinnGen data at the phenotype level. The median age of first event was used in the analysis. For disease traits from other resources, the age of onset was also looked up in FinnGen if the same disease exists in FinnGen. If a disease trait is not available in FinnGen, we looked it up from UpToDate (www.uptodate.com), which provides information on disease epidemiology. In general, we considered schizophrenia, eating disorder, type 1 diabetes, IBS, hayfever and some other respiratory diseases as early onset diseases, while age at first birth, PCOS and endometriosis are related to reproduction success. The details of the age of onset of diseases are shown in **Table S10**.

## Data Availability

All data produced in the present study are available upon reasonable request to the authors

## Author contribution

BB, NRW, AFM conceived the project. BB performed formal data analysis with input from AFM, VH and NRW. All authors contributed to the writing, editing and approval of the manuscript.

## Data and materials availability

FinnGen R6 GWAS summary statistics (https://finngen.gitbook.io/documentation/data-download). UK Biobank GWAS summary statistics round 2 (http://www.nealelab.is/uk-biobank). GWAS Catalog (https://www.ebi.ac.uk/gwas/)

